# Metabolic disturbances and inflammatory dysfunction predict severity of coronavirus disease 2019 (COVID-19): a retrospective study

**DOI:** 10.1101/2020.03.24.20042283

**Authors:** Shuke Nie, Xueqing Zhao, Kang Zhao, Zhaohui Zhang, Zhentao Zhang, Zhan Zhang

**Affiliations:** Department of Neurology, Renmin Hospital of Wuhan University, Wuhan 430060, China; Department of Respiratory Disease and Intensive Care, Renmin Hospital of Wuhan University, Wuhan 430060, China; Department of hepatology of traditional Chinese and Western medicine, the third people’s Hospital of Hubei province, Wuhan 430033, China

**Keywords:** Coronavirus disease 2019, infectious disease, metabolism, inflammation, severity prediction

## Abstract

**Background:** The coronavirus disease 2019 (COVID-19) is spreading worldwide with 16,558 deaths till date. Serum albumin, high-density lipoprotein (HDL-C), and C-reactive protein have been known to be associated with the severity and mortality of community-acquired pneumonia. However, the characteristics and role of metabolic and inflammatory indicators in COVID-19 is unclear.

**Methods:** We included 97 hospitalized patients with laboratory-confirmed COVID-19. Epidemiological, clinical, and laboratory indices; radiological features; and treatment were analysed. The differences in the clinical and laboratory parameters between mild and severe COVID-19 patients and the role of these indicators in severity prediction of COVID-19 were investigated.

**Results:** All were Wuhan residents with contact with confirmed COVID-19 cases. The median age was 39 years (IQR: 30–59). The most common presenting symptoms were fever (58.8%), cough (55.7%), and fatigue (33%). Other features were lymphopenia, impaired fasting glucose, hypoproteinaemia, hypoalbuminemia, low high-density lipoproteinemia. Decrease in lymphocyte count, serum total protein, serum albumin, high-density lipoprotein cholesterol (HDL-C), ApoA1, CD3^+^T%, and CD8^+^T% were found to be valuable in predicting the transition of COVID-19 from mild to severe illness. Chest computed tomography (CT) images showed that the absorption of bilateral lung lesions synchronized with the recovery of metabolic and inflammatory indicators.

**Conclusions:** Hypoproteinaemia, hypoalbuminemia, low high-density lipoproteinemia, and decreased ApoA1, CD3^+^T%, and CD8^+^T% could predict severity of COVID-19. Lymphocyte count, total serum protein, and HDL-C may be potentially useful for the evaluation of COVID-19.

## Introduction

In December 2019, a novel coronavirus from patients with pneumonia was reported in Wuhan, Hubei Province, China, and rapidly spread throughout the world[1 2]. As of 24 March, 2020, there were 38,3407 laboratory-confirmed cases worldwide, resulting in 16,558 deaths. The disease spectrum analysis of 44,415 patients diagnosed with COVID-19 by the Chinese Centre for Disease Control found that the mild type accounted for 81%, severe type for 14%, critical type for 5%, and the overall case-fatality rate (CFR) was 2.3%, but the fatality rate in critically ill patients was as high as 49% [3]. In 2003, SARS caused 774 deaths in 29 countries, and the CFR was about 10%. The number of deaths caused by COVID-19 was much higher than that by SARS. Although the total mortality rate of COVID-19 was lower than that of SARS, the mortality rate of critically ill patients with COVID-19 was much higher [4]. During the process of clinical diagnosis and treatment of patients with COVID-19, we found that some patients with mild disease quickly deteriorated or even died. Since the patients with mild disease account for the majority of COVID-19 patients, and the mortality rate is high in critically ill patients (49%), there is an urgent need to identify factors that can predict the transition of mild COVID-19 patients to critical patients.

The seventh edition of the COVID-19 diagnosis and treatment plan issued by the National Health Commission of China indicates that in adult patients with COVID-19, the progressive decrease in peripheral blood lymphocytes and the progressive increase in inflammatory factors IL-6 and C-reactive protein are early warning indicators for the progression of mild patients to severe and critical types. Our early reports demonstrate that cellular immunity, as indicated by the number of CD3^+^, CD4^+^, and CD8^+^T cells may be related to the severity of the disease in COVID-19[5]. However, cellular immunity and inflammatory factors are not routinely tested, and not all medical institutions can detect them. Therefore, in addition to the above indicators, there is an urgent need for a more simple and accessible technique to predict the potential of developing severe type of COVID-19 in patients with mild disease. We found that there were abnormalities in the levels of fasting blood glucose (FBG), serum total protein, albumin, and blood lipid metabolism of patients with COVID-19. In this study, we compared the levels of fasting blood glucose, serum total protein, albumin, blood lipid, cytokines, cellular and humoral immunity indices in patients with different severity types of COVID-19 and their clinical course. Furthermore, we studied the correlation between these indices and evaluated the role of biochemical metabolism and immune inflammation in predicting the development of severe COVID-19.

## Materials and Methods

### Study participants and design

This study was approved by the institutional ethics board of Renmin Hospital of Wuhan University. Requirement for written informed consent was waived by the ethics board of Renmin Hospital of Wuhan University (WDRY2020-K100). The patients with confirmed COVID-19 were admitted to Renmin Hospital from 9 February to 28 February, 2020. All patients with COVID-19 enrolled in this research study were laboratory-confirmed cases with positive results for fluorescence reverse transcription polymerase chain reaction (RT-PCR) detection of SARS-CoV-2. The final date of follow up was 10 March, 2020. According to the guidelines for diagnosis and treatment plan for COVID-19 issued by the National Health Commission of China[6], patients with COVID-19 were divided into three main types according to the clinical manifestations: (1) Mild: with fever, respiratory or digestive symptoms, and physician-diagnosed pneumonia by chest computed tomography (CT). (2) Severe: Having one of the following features: shortness of breath and respiratory rate (RR) ≥ 30 times/min; oxygen saturation ≤ 93% at rest; chest CT imaging showing that the lesion had progressed more than 50% within 24–48 hours. (3) Critical: Meeting any of the following criteria: respiratory failure needing mechanical ventilation; shock; and additional organ failure needing critical care and treatment. In response to the requirements of the Hubei provincial government with regard to treatment of COVID-19 patients according to grades, Renmin Hospital of Wuhan University only received mild and severe patients. Therefore, the patients included in this study were patients with mild and severe COVID-19. According to the course of the disease (from the onset of COVID-19-related symptoms to the testing time), we divided the mild patients into two groups: the course of disease in group one was 14 days (mild 1 group), and in the other group was 30 days (mild 2 group). The average course of disease in severe patients (severe group) was 12 days.

### Laboratory Confirmation and Detection

Nasopharyngeal and anal swab samples were collected and fluorescence RT-PCR was used to detect open reading frame 1ab (open reading frame1ab, ORF1ab) and nucleocapsid protein (nucleocapsid protein, N) of 2019-nCoV. RT-PCR procedure was performed according to the protocol issued by the World health Organisation (WHO). The novel coronavirus (2019-nCoV) ORF1ab/N gene double nucleic acid detection kit (fluorescent PCR method) was purchased from the Shanghai Jienuo Company. The detection equipment QuantstudioDx and 7500 fluorescent PCR instrument were purchased from ThermoFisher Company of the United States. The 2019-nCov-positive case confirmation needs to meet the requirement that both the target genes are positive simultaneously, or that the ORF1ab is positive in two different samples from the same patient.

To investigate the effect of 2019-nCoV on the production of cytokines and immunity in different types and stages of the illness, plasma cytokines (IL-2, IL-4, IL-5, IL-6, IL-10, tumor necrosis factor, and interferon-γ), cellular immunity indicators (CD3^+^, CD4^+^, CD8^+^, CD19^+^, CD16^+^56), and humoral immunity indicators (Immunoglobulin G [IgG], Immunoglobulin M [IgM], Immunoglobulin A [IgA], Immunoglobulin E [IgE], Complement C3, Complement C4) were measured according to the manufacturer’s protocol.

### Data Collection

We obtained the medical records of patients with COVID-19, as reported to the National Health Commission of China. Data on blood routine and fasting blood glucose, liver function, renal function, total protein and albumin, full set of lipids, CRP, creatine kinase, lactate dehydrogenase, cytokines, humoral immunity, and cellular immunity indicators were extracted. The data were analysed by the research team of Renmin Hospital of Wuhan University. Exposure history, clinical symptoms, laboratory tests data, radiological features, and treatment data were obtained from the medical records from 9 February to February 28, 2020.

### Statistical Analysis

Categorical variables were described as frequency rates and percentages, and continuous variables were described using median and interquartile range (IQR) values. For numerical variables, we first performed the Gaussian distribution test. Means for continuous variables were compared using independent sample *t* tests when data were normally distributed; otherwise, nonparametric Wilcoxon rank sum test was used. For categorical variables, the comparison between groups was made by χ2 test, and the Fisher exact test was used when the data were limited. Hotmap of correlation analysis was performed by Graphpad Prism 8. To evaluate the value of metabolic and inflammatory indices in predicting the severity of COVID-19, a receiver-operating characteristics (ROC) curve and the area under the ROC curve (AUROC) were performed. All statistical analyses were performed using SPSS 26 software. *P* value less than 0.05 was considered statistically significant.

## Results

### Demographic and Clinical Characteristics of Patients with COVID-19

This study included 97 hospitalized patients with confirmed COVID-19. The median age of the patients was 39 years (interquartile 30–60; range 23–82) and 63 (64.9%) patients were women. All the patients were residents of Wuhan without contact with wildlife, but all had contact with patients with COVID-19. Compared with patients in the mild group (n = 72), the patients in the severe group (n = 25) were significantly older (median age, 58 years [IQR, 47–67] vs. 37 years [IQR, 29–55]; *p* <0.001) and were more likely to have clinical comorbidities, including hypertension (10 [40%] vs. 5 [6.9%]), diabetes (2 [8%] vs. 3 [4.2%]), cardiovascular disease (2 [8%] vs. 0 [0%]), and cerebrovascular disease (2 [8%] vs. 1 [1.4%]) (Table 1).

**Table 1.** Demographic and clinical characteristics of patients infected with COVID-19.

|  | No.( %) |  |  | *p* value |
| --- | --- | --- | --- | --- |
|  | Total<br>(n = 97) | Mild<br>(n = 72) | Severe (n =<br>25) |  |
| **Age, median(IQR), years** | 39(30-60) | 37(29-55) | 58(47-67) | **<0.001** |
| **Sex** |  |  |  | 0.42 |
| Men | 34(35.0) | 21(29.1) | 13(52.0) |  |
| Women | 63(64.9) | 51(70.8) | 12(48.0) |  |
| **Any comorbidity** |  |  |  |  |
| Hypertension | 15(15.5) | 5(6.9) | 10(40.0) | **<0.001** |
| Diabetes | 5(5.2) | 3(4.2) | 2(8.0) | 0.475 |
| Hyperlipidemia | 7(7.2) | 3(4.2) | 4(16.0) | 0.067 |
| Chronic pulmonary disease | 2(2.1) | 1(1.4) | 1(4.0) | 0.459 |
| Coronary heart disease | 2(2.1) | 0 | 2(8.0) | **0.019** |
| Cerebrovascular atherosclerosis | 3(3.1) | 1(1.4) | 2(8.0) | 0.131 |
| Malignancy | 3(3.1) | 1(1.4) | 2(8.0) | 0.131 |
| Chronic kidney disease | 3(3.1) | 1(1.4) | 2(8.0) | 0.131 |
| Chronic liver disease | 3(3.1) | 0 | 3(12.0) | **0.004** |
| **Signs and symptoms** |  |  |  |  |
| Asymptomatic | 7(7.2) | 7(9.7) | 0 | **0.037** |
| Fever | 57(58.8) | 36(50.0) | 21(84.0) | **0.002** |
| Cough | 54(55.7) | 36(50.0) | 18(72.0) | 0.053 |
| Sputum production | 15(15.5) | 9(12.5) | 6(24.0) | 0.187 |
| Myalgia or fatigue | 32(33.0) | 29(40.3) | 3(12.0) | **0.006** |
| Nausea or Vomit | 5(5.2) | 1(1.4) | 4(16.0) | **0.009** |
| Diarrhea | 12(12.4) | 8(11.1) | 4(16.0) | 0.532 |
| Sore throat | 10(10.3) | 7(9.7) | 3(12.0) | 0.75 |
| Dyspnea | 8(8.2) | 3(4.2) | 5(20.0) | **0.022** |
| Dizzy and Headache | 7(7.2) | 5(6.9) | 2(8.0) | 0.862 |
| Itchy eyes | 1(1.0) | 1(1.4) | 0 | 0.439 |

Of the 97 patients in this study, asymptomatic cases accounted for 7.2% of the patients. Analysis of the clinical characteristics of 72,314 COVID-19 patients by the Chinese Centre for Disease Control revealed that asymptomatic cases accounted for 1%[3]. The most common symptoms at onset of illness were fever (58.8% on admission), cough (55.7%), and fatigue (33%). The second most common symptoms were sputum production (15.5%), vomiting and diarrhoea (12.4%), nasal congestion (10.3%), and neurological symptoms such as dizziness and headache (7.2%) (Table 1).

### Laboratory Parameters in Patients with COVID-19

According to the course of disease, the patients with mild type COVID-19 were divided into two groups: the average course of disease was 14 days in the mild 1 group and 30 days in mild 2 group. Lymphopenia was a feature of patients with severe COVID-19, and a lower lymphocyte count was found in the severe group compared with the mild 1 group (0.99[0.78–1.43] vs. 1.9[1.4–2.3]). The levels of apolipoproteins A1(ApoA1) (1.52[1.40–1.60] vs. 1.37[1.23–1.59]) and ApoB (0.88[0.71–1.05] vs. 0.76[0.66–0.87]) were significantly increased in mild 2 group compared with mild 1 group. There was no statistical difference observed in the hepatic and renal function between the two groups of patients with mild COVID-19. The pattern of impaired fasting blood glucose (FBG) in mild COVID-19 patients was fasting hypoglycaemia, while that in severe patients was predominantly fasting hyperglycaemia. Fasting hypoglycaemia was found in 21.4% of patients in mild 1 group with no case of fasting hyperglycaemia. In the mild 2 group, 34.1% of the patients had fasting hypoglycaemia, and 2.3% had fasting hyperglycaemia. Compared with mild COVID-19 patients, we found that 24% of severe COVID-19 patients had fasting hyperglycaemia and 4% had fasting hypoglycaemia. Patients in the severe group had a lower level of serum total protein (59[58–63] vs. 65[63–70]), serum albumin (36[34–39] vs. 41[37–43]), total cholesterol (3.6[3.3–4.0] vs. 3.8[3.5–4.4]), and HDL-C (0.88[0.81–1.10] vs. 1.05[0.93–1.50]) compared with the mild 1 group (Table 2).

**Table 2.**
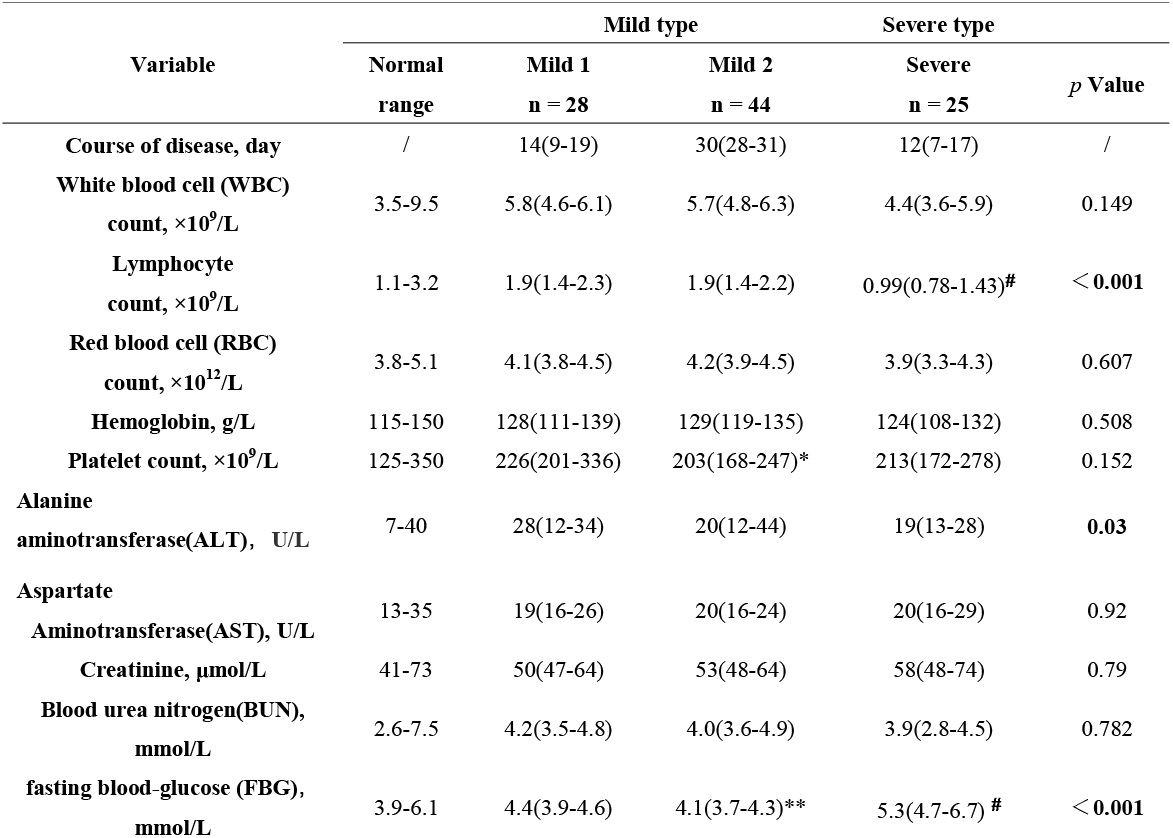
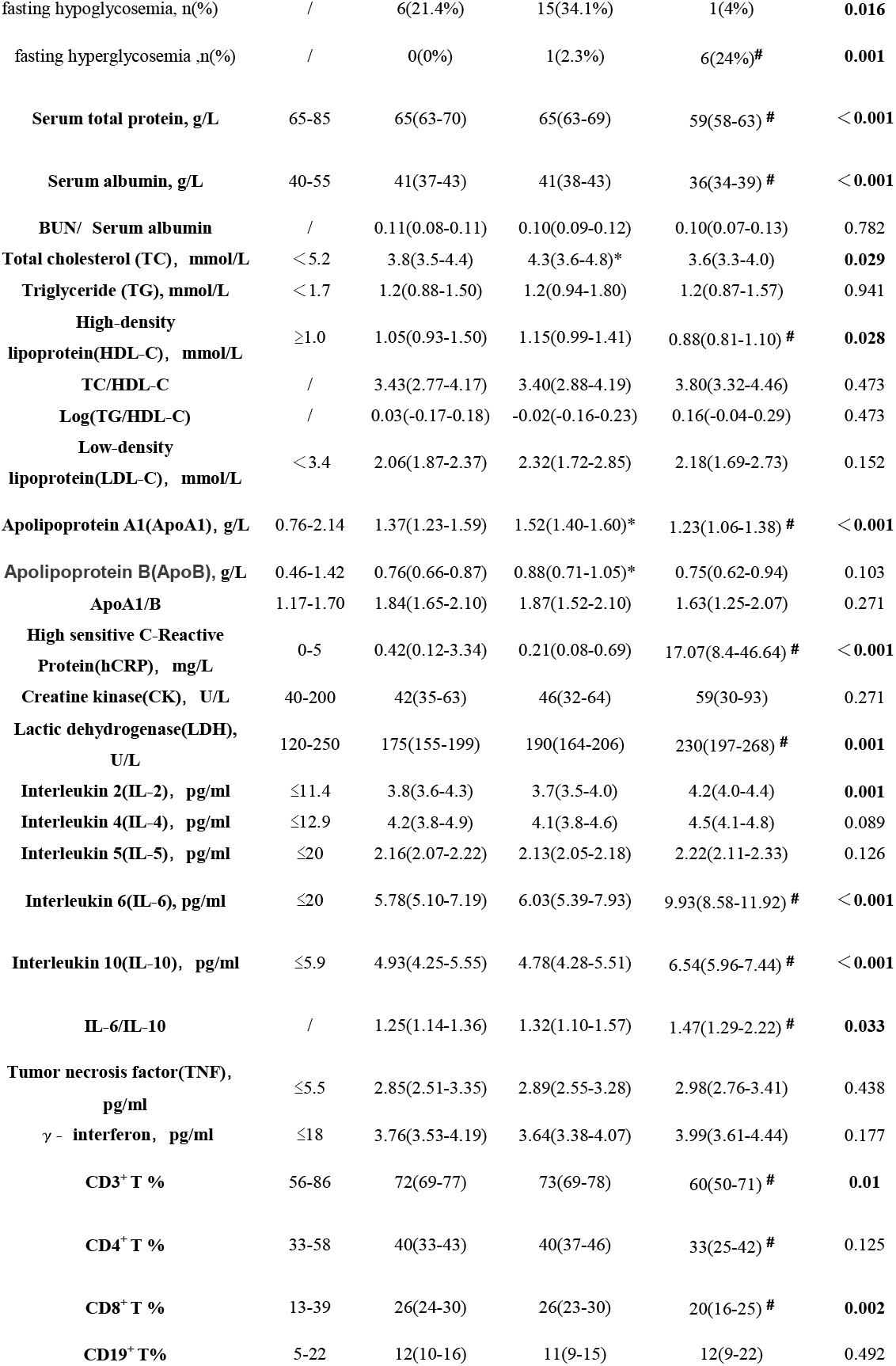
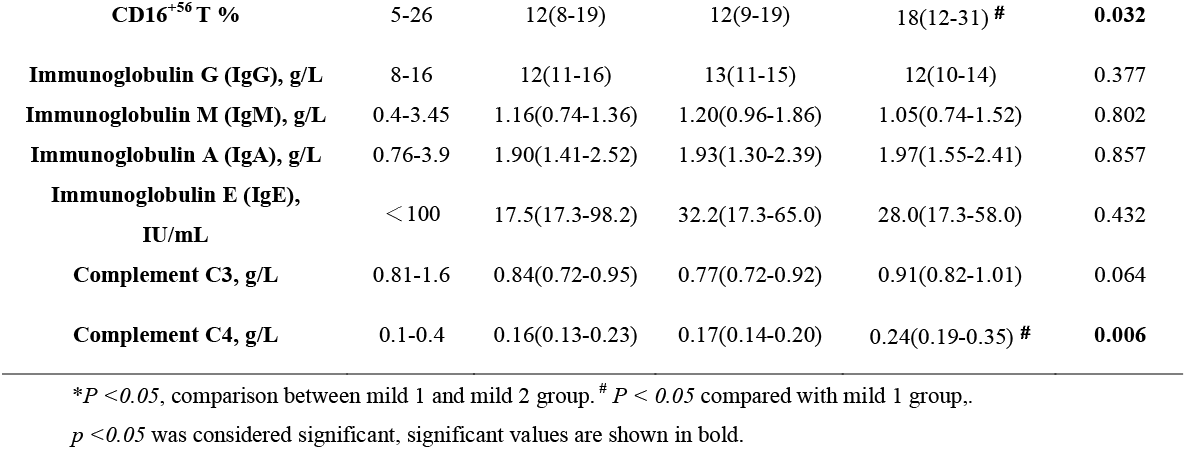
Laboratory Findings of Patients with COVID-19 during hospitalization (Median (IQR))

The levels of serum lactate dehydrogenase (230[197–268)] vs. 175[155–199]), IL-2 (4.2[4.0–4.4] vs. 3.8[3.6–4.3]), IL-6 (9.93[8.58–11.92] vs. 5.78[5.10–7.19], and IL-10 (6.54[5.96–7.44] vs. 4.93[4.25–5.55]) in severe patients were significantly higher than those in mild patients with COVID-19, suggesting an overactive inflammatory response in severe illness. Furthermore, compared to the mild group, the proportion of CD3^+^T cells (60%[50%–71%] vs. 72%[69%–77%]) and CD8^+^T cells (20%[16%–25%] vs. 26%[24%–30%]) was significantly decreased, while the proportion of CD16^+^56T cells was increased (18[12–31] vs. 12[8–19]), in the severe group indicating impaired cellular immunity. The level of complement C4 in patients with severe COVID-19 was higher than that in the mild group (0.24[0.19–0.35] vs. 0.16[0.13–0.23]) (Table 2).

### Relationship between the metabolic and inflammatory indices in patients with COVID-19

In order to investigate the correlation between metabolic abnormalities and severity of disease in COVID-19, partial correlation analysis was performed on the basis of controlling the variables of CRP, inflammatory factors (IL-2, IL-6, IL-10, IL-6/IL-10), and cellular immunity indicators (CD3^+^, CD8^+^, CD16^+^56), C3 and C4 (Figure 1). It was found that the severity of COVID-19 positively correlated with FBG (r = 0.334, *p* = 0.023). Serum total protein (r = −0.422, *p* = 0.004), serum albumin (r = −0.351, *p* = 0.017), HDL-C (r = −0.332, *p* = 0.024), and ApoA1 (r = −0.325, *p* = 0.028) displayed negative correlation with the severity of COVID-19. FBG in patients with COVID-19 positively correlated with the level of CRP (r = 0.353, *p*< 0.001), and negatively correlated with the lymphocyte count (r = 0.27, *p* = 0.008) and percentage of CD3^+^T cells (r = 0.206, *p* = 0.044). Serum total protein showed positive correlation with the percentage of CD8^+^T cells (r = 0.208, *p* = 0.042). Serum albumin displayed positive correlation with the lymphocyte count (r = 0.452, *p*<0.001) and CD8^+^T cell percentage (r = 0.237, *p* = 0.02), and negative correlation with the levels of CRP (r = −0.252, *p* = 0.013), IL-6 (r = −0.334, *p* = 0.001), IL-10 (r = −0.265, *p* = 0.009), and IL-6/IL-10 (r = −0.284, *p* = 0.005). Total cholesterol positively correlated with the percentage of CD3^+^T cells (r = 0.277, *p* = 0.006) and negatively correlated with the level of IL-6 (r = −0.219, *p* = 0.032). The level of HDL-C negatively correlated with the level of IL-2 (r = −0.222, *p* = 0.030) and IL-10 (r = −0.211, *p* = 0.039) in patients with COVID-19, and ApoA1 negatively correlated with the level of CRP (r = −0.371, *p*<0.001), IL-2 (r = −0.207, *p* = 0.043), IL-6 (r = −0.237, *p* = 0.020), and IL-10 (r = −0.252, *p* = 0.013) (Table 3).

**Table 3.**
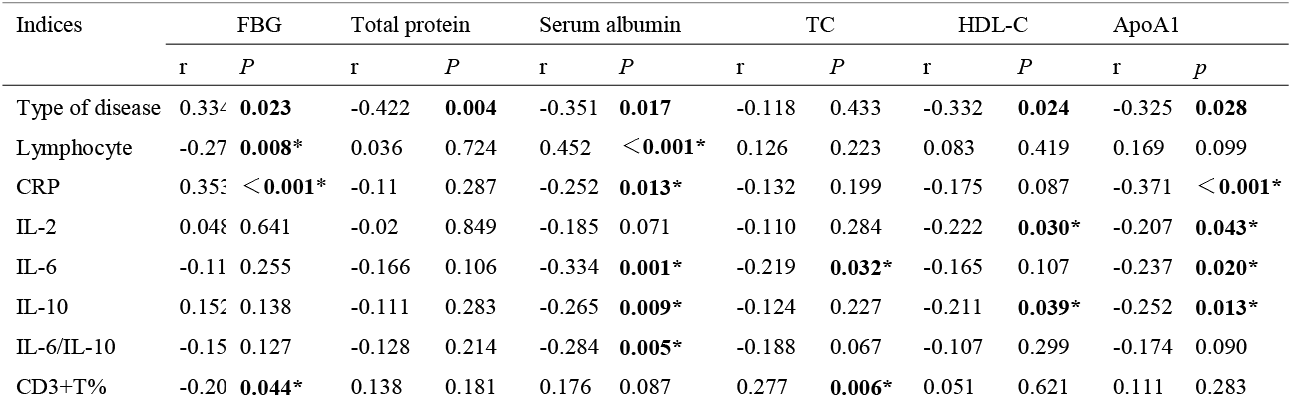
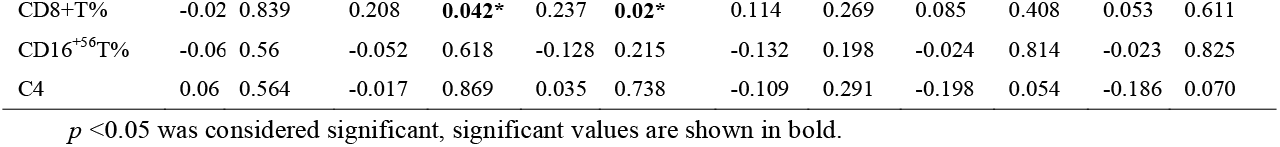
Correlation between metabolic indicators and inflammatory in patients with COVID-19.

| Indices | FBG |  | Total protein |  | Serum albumin |  | TC |  | HDL-C |  | ApoA1 |  |
| --- | --- | --- | --- | --- | --- | --- | --- | --- | --- | --- | --- | --- |
|  | r | P | r | P | r | P | r | P | r | P | r | p |
| Type of disease | 0.334 | **0.023** | -0.422 | **0.004** | -0.351 | **0.017** | -0.118 | 0.433 | -0.332 | **0.024** | -0.325 | **0.028** |
| Lymphocyte | -0.27 | **0.008\*** | 0.036 | 0.724 | 0.452 | **<0.001\*** | 0.126 | 0.223 | 0.083 | 0.419 | 0.169 | 0.099 |
| CRP | 0.353 | **<0.001\*** | -0.11 | 0.287 | -0.252 | **0.013\*** | -0.132 | 0.199 | -0.175 | 0.087 | -0.371 | **<0.001\*** |
| IL-2 | 0.048 | 0.641 | -0.02 | 0.849 | -0.185 | 0.071 | -0.110 | 0.284 | -0.222 | **0.030\*** | -0.207 | **0.043\*** |
| IL-6 | -0.11 | 0.255 | -0.166 | 0.106 | -0.334 | **0.001\*** | -0.219 | **0.032\*** | -0.165 | 0.107 | -0.237 | **0.020\*** |
| IL-10 | 0.152 | 0.138 | -0.111 | 0.283 | -0.265 | **0.009\*** | -0.124 | 0.227 | -0.211 | **0.039\*** | -0.252 | **0.013\*** |
| IL-6/IL-10 | -0.15 | 0.127 | -0.128 | 0.214 | -0.284 | **0.005\*** | -0.188 | 0.067 | -0.107 | 0.299 | -0.174 | 0.090 |
| CD3+T% | -0.20 | **0.044\*** | 0.138 | 0.181 | 0.176 | 0.087 | 0.277 | **0.006\*** | 0.051 | 0.621 | 0.111 | 0.283 |
| CD8+T% | -0.02 | 0.839 | 0.208 | **0.042\*** | 0.237 | **0.02\*** | 0.114 | 0.269 | 0.085 | 0.408 | 0.053 | 0.611 |
| CD16<sup>+</sup>56T% | -0.06 | 0.56 | -0.052 | 0.618 | -0.128 | 0.215 | -0.132 | 0.198 | -0.024 | 0.814 | -0.023 | 0.825 |
| C4 | 0.06 | 0.564 | -0.017 | 0.869 | 0.035 | 0.738 | -0.109 | 0.291 | -0.198 | 0.054 | -0.186 | 0.070 |

$p < 0.05$ was considered significant, significant values are shown in bold.

**Figure 1.**
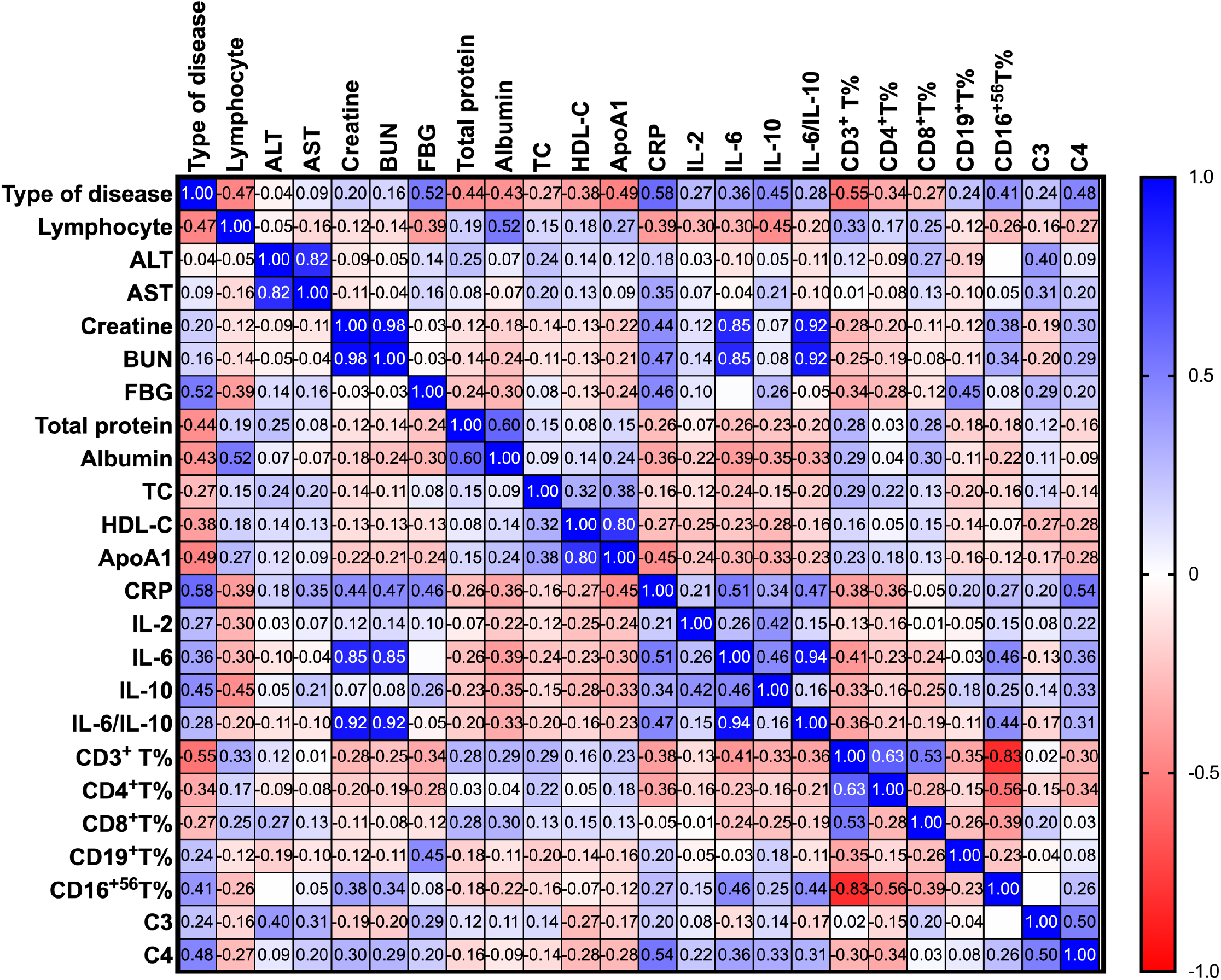
Correlation between the type of COVID-19 and laboratory parameters. The severity of COVID-19 showed positive correlation with FBG, and negative correlation with the levels of serum total protein, serum albumin, HDL-C, and ApoA1. The levels of creatine and blood urea nitrogen (BUN) displayed significant positive correlation with IL-6 and IL-6/IL-10. The serum total protein level showed positive correlation with the percentage of CD8^+^T cells. Serum albumin concentration demonstrated positive correlation with the lymphocyte count and CD8^+^T%, and negative correlation with the levels of CRP, IL-6, IL-10, and IL-6/IL-10. Total cholesterol showed positive correlation with the percentage of CD3^+^T cells and negative correlation with the IL-6 level.

### Role of metabolic and inflammatory indicators in predicting severity of COVID-19

In order to evaluate the value of the metabolic and inflammatory indices for predicting severity of the disease, a receiver-operating characteristics (ROC) curve and the area under ROC curve (AUROC) were analysed. Decrease in lymphocyte count, serum total protein, serum albumin, HDL-C, ApoA1, CD3^+^T%, and CD8^+^T% were of considerable value in predicting the transition of COVID-19 from mild to severe illness. Among the above indices, lymphocyte count and serum total protein had more predictive value. AUROC and 95% confidence interval (95% CI) of lymphocyte count, serum total protein, serum albumin, and CD3^+^T% were 0.838 (95% CI: 0.727–0.949), 0.833 (95% CI: 0.714–0.951), 0.781 (95% CI: 0.652–0.910) and 0.784 (95% CI: 0.656–0.913), respectively (Figure 2). The severity predictive value of lymphopenia was improved by adding serum total protein. AUROC of lymphocyte count plus total protein was 0.899 (95% CI: 0.811–0.986) (Table 4).

**Table 4.** Analysis of metabolic and inflammatory indicators in predicting severity of Covid-19.

|  | Mild 1 group | Sever group | AUROC (95%CI) | *p* |
| --- | --- | --- | --- | --- |
|  | Median (IQR) | Median (IQR) |  |  |
| **Lymphocyte** | 1.9 (1.4-2.3) | 0.99(0.78-1.43) | 0.838 (0.727-0.949) | **<0.001** |
| **Total protein** | 65(63-70) | 59(58-63) | 0.833 (0.714-0.951) | **<0.001** |
| **Lymphocyte + total protein** | / | / | 0.899(0.811-0.986) | **<0.001** |
| **Albumin** | 41(37-43) | 36(34-39) | 0.781 (0.652-0.910) | **<0.001** |
| **HDL-C** | 1.05(0.93-1.50) | 0.88(0.81-1.10) | 0.707 (0.566-0.848) | **0.010** |
| **ApoA1** | 1.37(1.23-1.59) | 1.23(1.06-1.38) | 0.728 (0.592-0.864) | **0.004** |
| **CD3<sup>+</sup> T%** | 72(69-77) | 60(50-71) | 0.784 (0.656-0.913) | **<0.001** |
| **CD8<sup>+</sup> T%** | 26(24-30) | 20(16-25) | 0.720 (0.573-0.867) | **0.006** |

**Figure 2.**
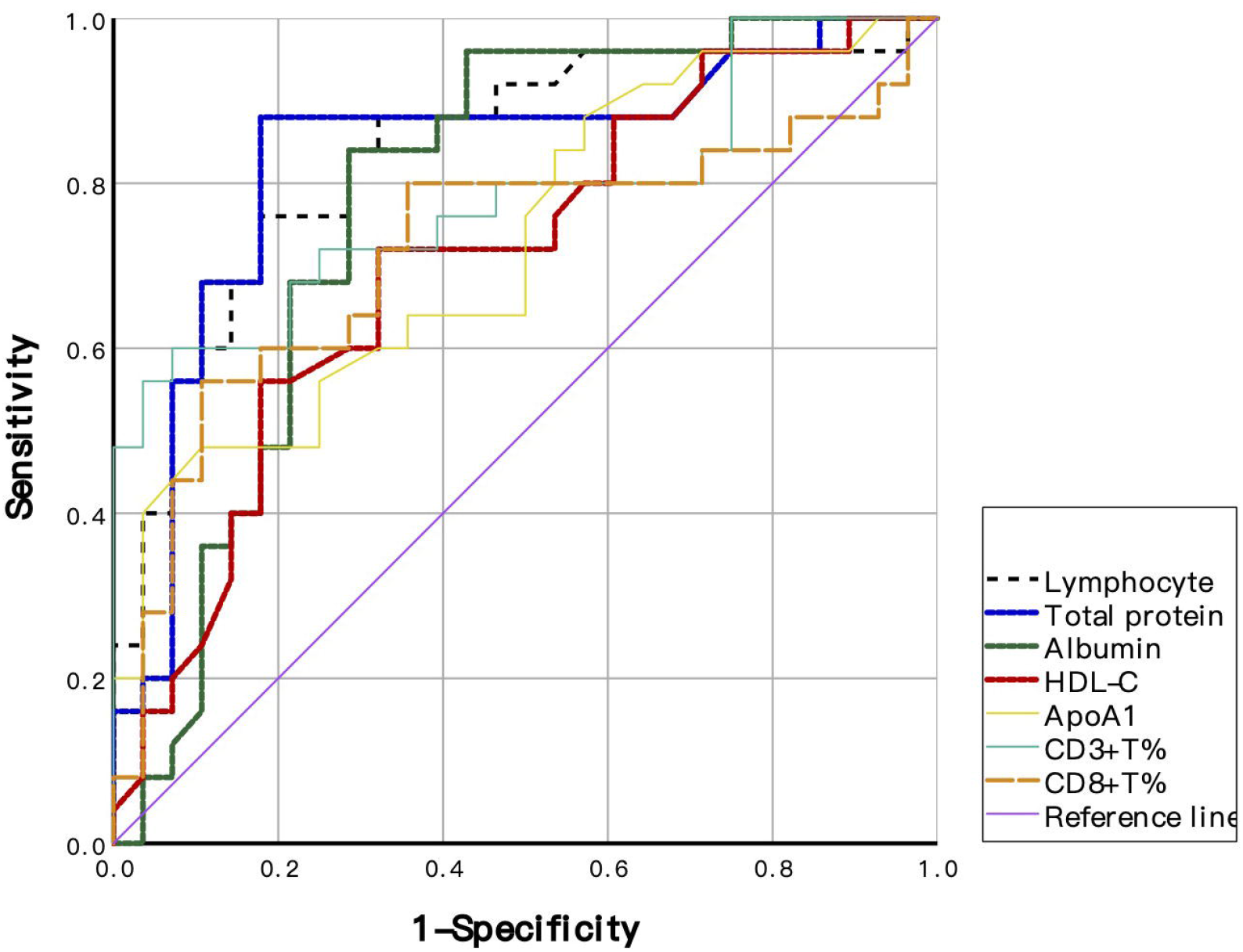
Analysis of the receiver-operating characteristics (ROC) curve for predicting the severity of COVID-19. Decrease in lymphocyte count, serum total protein, serum albumin, HDL-C, ApoA1, CD3^+^T%, and CD8^+^T% were of great value in predicting the transition of COVID-19 from mild to severe illness. Among the above indices, lymphocyte count and serum total protein had more predictive value.

### Treatments and outcomes of patients with COVID-19

All patients in this study were treated with bed rest and supportive treatment. A total of 86 (88.7%) patients received antiviral therapy (oseltamivir or arbidol), 47 (48.5%) received antibiotic therapy, 35 (36.1%) received immunomodulatory therapy (hydroxychloroquine or chloroquine phosphate), 27 (27.8%) patients were given short-term (3–5 days) and low-dose systematic corticosteroids. Higher percentages of severe patients received these therapies (Table 5). Nasal cannula oxygen support was given to 15 patients (15.5%), with a higher percentage among the severe patients than patients in the mild 1 group (40% vs. 7.1%). As of 10 March, 2020, which was the final date of follow up, out of the 97 patients, 66 (68%) had been discharged, 30 (30.9%) were still hospitalized, and one (1%) had died. Severe patients with COVID-19 had a higher chance of hospitalization than those with mild disease (52% vs. 28.6%) (Table 5).

**Table 5.**
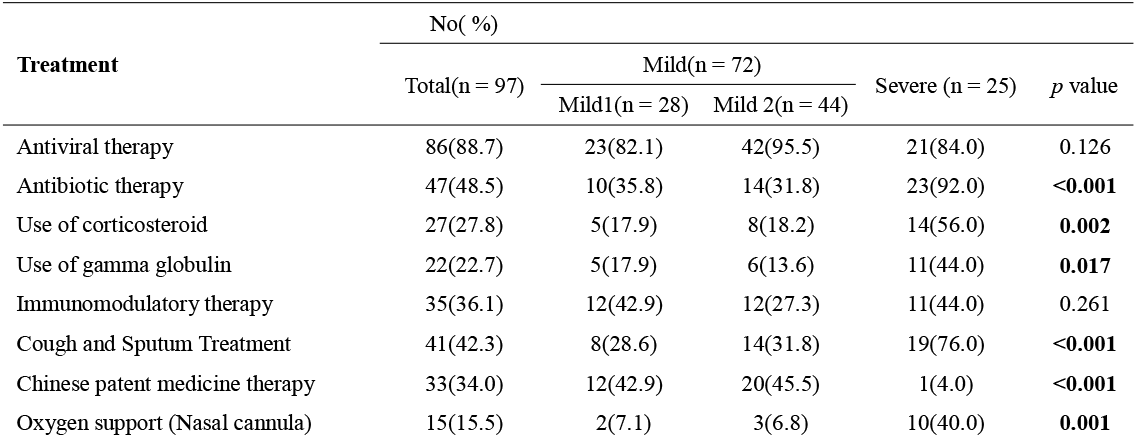
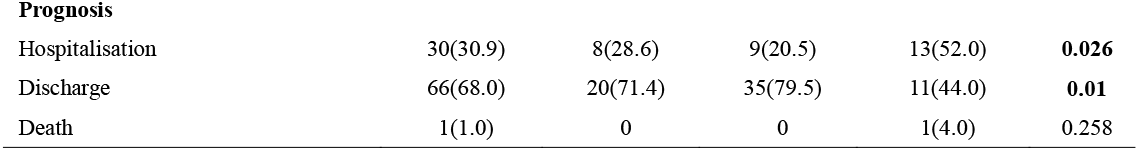
Treatments and outcomes of patients with COVID-19.

### Dynamic profile of laboratory tests in patients with COVID-19

To investigate the major laboratory and radiological features that appeared during COVID-19 progression, the dynamic changes in seven laboratory parameters and chest CT of two severe patients were evaluated (Table 6). By 6 March, 2020, both the patients had been discharged with negative novel coronavirus RNA nasopharyngeal swab test and absorbed lesions on CT imaging. During hospitalization, majority of the patients had lymphopenia, hypoproteinaemia, hypoalbuminemia, and low high-density lipoproteinemia in the early stage of the disease. However, severe COVID-19 patients showed worsening of the parameters over time. Additionally, we observed that the red blood cell count and haemoglobin concentration progressively decreased during COVID-19 progression.

**Table 6.**
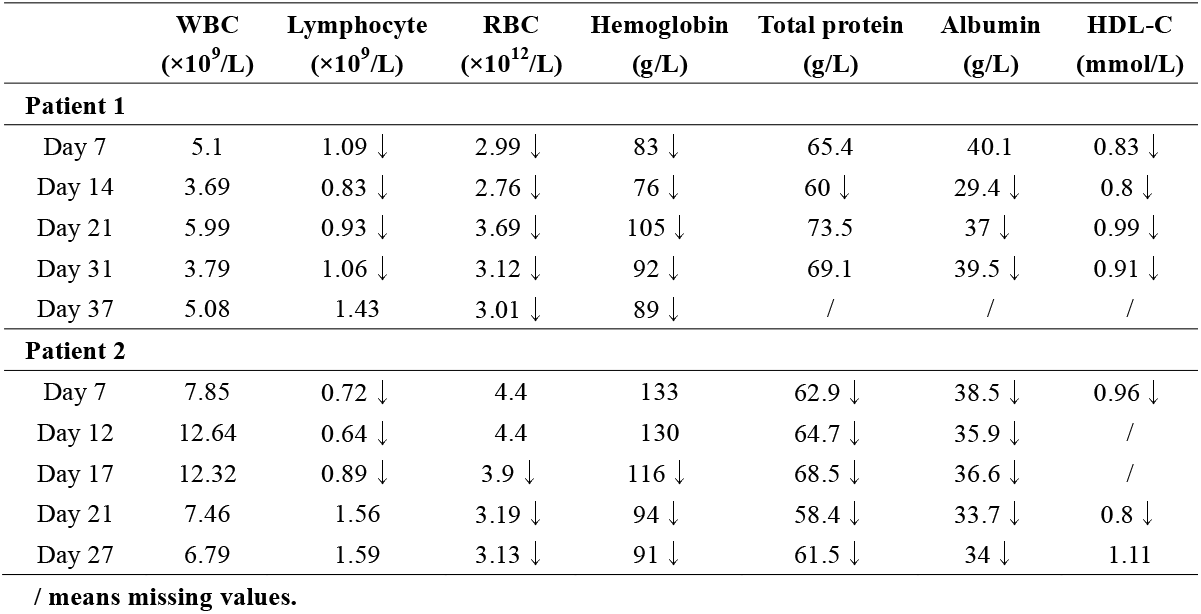
Dynamic Profiles of Laboratory Parameters in two severe patients with COVID-19.

| | **WBC**<br>($\times 10^9/L$) | **Lymphocyte**<br>($\times 10^9/L$) | **RBC**<br>($\times 10^{12}/L$) | **Hemoglobin**<br>(g/L) | **Total protein**<br>(g/L) | **Albumin**<br>(g/L) | **HDL-C**<br>(mmol/L) |
| --- | --- | --- | --- | --- | --- | --- | --- |
| **Patient 1** |  |  |  |  |  |  |  |
| Day 7 | 5.1 | 1.09 ↓ | 2.99 ↓ | 83 ↓ | 65.4 | 40.1 | 0.83 ↓ |
| Day 14 | 3.69 | 0.83 ↓ | 2.76 ↓ | 76 ↓ | 60 ↓ | 29.4 ↓ | 0.8 ↓ |
| Day 21 | 5.99 | 0.93 ↓ | 3.69 ↓ | 105 ↓ | 73.5 | 37 ↓ | 0.99 ↓ |
| Day 31 | 3.79 | 1.06 ↓ | 3.12 ↓ | 92 ↓ | 69.1 | 39.5 ↓ | 0.91 ↓ |
| Day 37 | 5.08 | 1.43 | 3.01 ↓ | 89 ↓ | / | / | / |
| **Patient 2** |  |  |  |  |  |  |  |
| Day 7 | 7.85 | 0.72 ↓ | 4.4 | 133 | 62.9 ↓ | 38.5 ↓ | 0.96 ↓ |
| Day 12 | 12.64 | 0.64 ↓ | 4.4 | 130 | 64.7 ↓ | 35.9 ↓ | / |
| Day 17 | 12.32 | 0.89 ↓ | 3.9 ↓ | 116 ↓ | 68.5 ↓ | 36.6 ↓ | / |
| Day 21 | 7.46 | 1.56 | 3.19 ↓ | 94 ↓ | 58.4 ↓ | 33.7 ↓ | 0.8 ↓ |
| Day 27 | 6.79 | 1.59 | 3.13 ↓ | 91 ↓ | 61.5 ↓ | 34 ↓ | 1.11 |

/ means missing values.

Chest CT scan demonstrated bilateral multiple small patches and interstitial changes in the lungs after symptom onset (Figure 3). The lesions seen on chest CT gradually increased and reached a peak with progressive lymphopenia, hypoproteinaemia, hypoalbuminemia, and low high-density lipoproteinemia (Figure 3 and Table 6). In the late stage of COVID-19, absorption of bilateral ground glass lesions was observed following treatment along with the recovery of lymphocyte count, and increase in total protein, albumin, and HDL-C.

**Figure 3.**
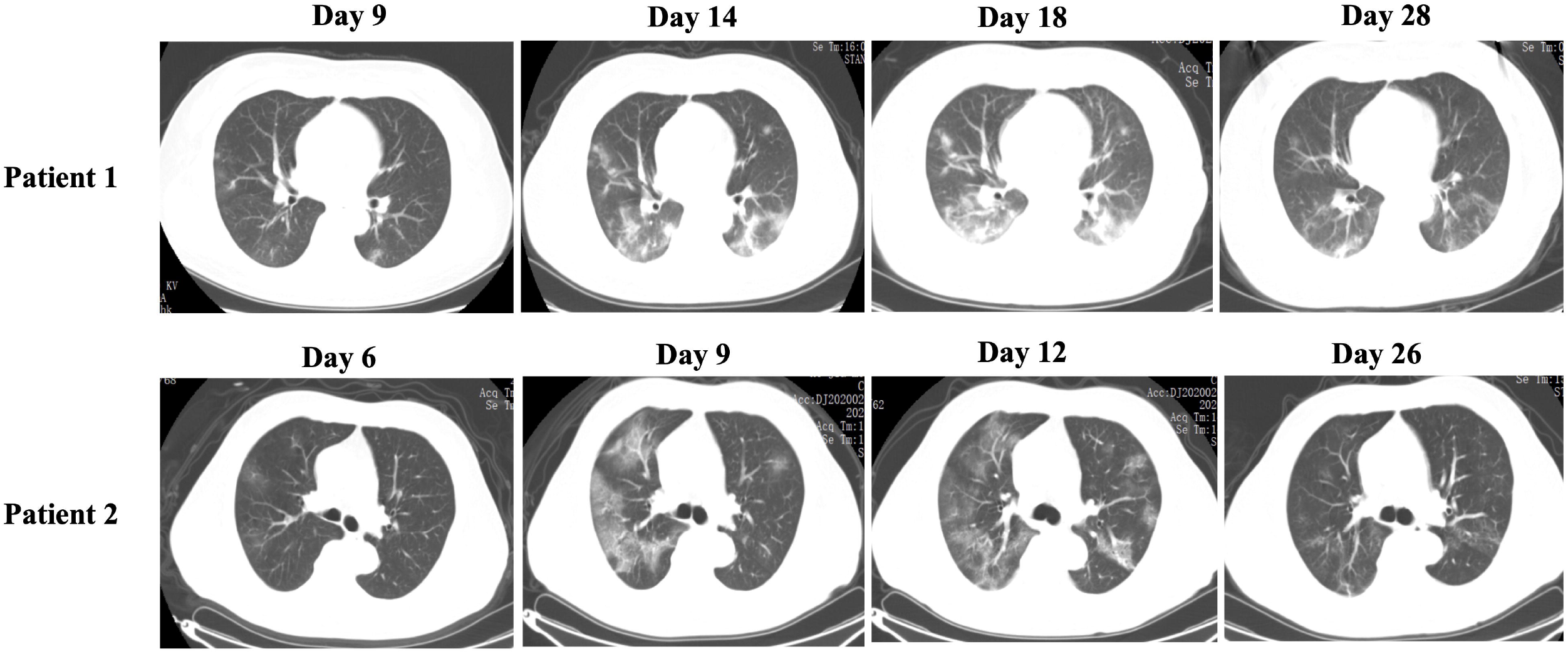
Dynamic profiles of chest computed tomographic images of two patients with severe COVID-19 after symptom onset. Chest CT scans revealed bilateral patches and ground-glass opacities in the lungs, that gradually increased and reached a peak, followed by absorption with treatment and recovery of metabolic and inflammatory indicators.

## Discussion

The consequences of human exposure to the SARS-CoV-2 virus are closely related to the functional status of the body, in addition to the virulence and dose of the exposure. Looking for early warning indicators for severe type of COVID-19 may be helpful to reduce the mortality. We found that the combination of a variety of chronic diseases was one of the factors that contributed to high mortality in critically ill patients with COVID-19. The coexisting disorders in patients with severe COVID-19 in our study included hypertension (40%), cardiovascular disease (8%), and diabetes (8%). A higher percentage of comorbidities (hypertension and coronary heart disease) was found in severe patients than in mild patients with COVID-19. An analysis of the clinical characteristics of 1,099 patients with COVID-19 revealed that 23.7% of the patients had coexisting diseases, including hypertension (15%), diabetes (7.4%), coronary heart disease (2.5%), cerebrovascular disease (1.4%), and chronic kidney disease (0.7%)[7]. Out of the above 1,099 patients, 15.7% had the critical type of COVID-19, and 38.7% had other chronic diseases, including hypertension (23.7%), diabetes (16.2%), coronary heart disease (5.8%), cerebrovascular disease (2.3%), and chronic kidney disease (1.7%). CFR of the patients with comorbidities was significantly higher than that of patients without comorbidities[3]. Cardiovascular diseases, especially hypertension, may increase the risk of illness and death from COVID-19. SARS-CoV-2 infection might attack the cardiovascular system, leading to acute myocardial injury, hypotension, and tachycardia[8]. The specific role of cardiovascular system in the pathogenesis of COVID-19 is still unclear. The high expression of angiotensin converting enzyme 2 (ACE2) in the endothelial cells of the coronary arteries and the immune damage caused by the cytokines released during the inflammatory storms in critically ill patients might explain the involvement of the cardiovascular system [8 9].

The manifestation of impaired FBG in patients with mild COVID-19 was fasting hypoglycaemia, while that in severe patients was predominantly fasting hyperglycaemia. Since most of these patients did not have a history of diabetes, this might indicate impaired glucose regulation in the pathogenesis of COVID-19 with severe COVID-19 patients consuming more energy. These results suggest that supportive care and maintenance of caloric intake are crucial during treatment. Progressive decrease in blood lymphocytes and increase in IL-6 and CRP were found to be early clinical warning indicators for the progression of mild COVID-19 patients to severe and critical patients, which were consistent in our study. Moreover, we demonstrated that serum total protein, serum albumin, HDL-C, ApoA1, CD3^+^T%, and CD8^+^T% were of significant value in predicting the progression of patients with mild COVID-19 to severe and critical types of COVID-19. Serum albumin has been thought to be an independent risk factor for mortality in patients with community-acquired infectious diseases[10]. Hypoproteinaemia and hypoalbuminemia are indicative of malnutrition and underlying infectious processes. In our study, the predictive value of lymphocyte count for predicting the severity of COVID-19 was improved by adding serum total protein. HDL-C and ApoA1 are known to play protective roles in normal health and function of the lungs, as well as in a variety of disease states, including viral pneumonia[11]. Serum HDL-C levels and serum albumin levels might decrease and serum total cholesterol/HDL-C ratios and log (TG/HDL-C) values might increase proportionally in community-acquired pneumonia[12]. In our study, hypoproteinaemia, hypoalbuminemia, and low high-density lipoproteinemia were discovered in both mild and severe patients with COVID-19, but were much worse in severely ill patients, thus suggesting that metabolic disturbances contributed to the development of COVID-19. It has been reported that blood urea nitrogen (BUN) to serum albumin ratio could be used as a prognostic factor for mortality in patients with aspiration pneumonia [13]. However, this was not confirmed in our study. Liver function damage was found to be more frequent in COVID-19 patients than in non-COVID-19 patients[14]. In the present study, liver and kidney dysfunction were not apparent in patients with COVID-19, possibly because only mild and severe cases of COVID-19 were included in this study and liver and renal function damage mainly occurs in critically ill patients.

IL-10 is a very important anti-inflammatory cytokine in pneumonia. In our study, the level of IL-10 was higher in severe patients with COVID-19 than in mild patients. The levels of IL-6, IL-10, and IL-6/IL-10 ratio were significantly elevated in the severe group, suggesting the development of an overactive inflammatory response during the progression of the disease. A recent pathological study demonstrated that acute respiratory distress syndrome (ARDS) was the common immunopathological event in SARS-CoV-2 and SARS-CoV infections[15]. Cytokine storm is a feature of ARDS, and a severe systemic inflammatory response resulting from the release of large amounts of pro-inflammatory cytokines (IFN-γ, IL-6, TNF-α, TGFβ, etc.) by immune effector cells develops in SARS-CoV infection, which might also occur in SARS-CoV-2 infection[16]. Our previous study revealed that the number of cellular immunity CD3^+^, CD4^+^, and CD8^+^T cells may be related to the severity of COVID-19[5]. It has been shown that CD8^+^ T cells protect against pneumovirus-induced disease in mice[17]. In our study, the proportion of CD3^+^, CD8^+^, and CD16^+56^ T cells in the peripheral blood was significantly reduced in severe COVID-19 patients compared with mild patients, suggesting that these indices may be useful as an indicator of the disease status. Analysis of ROC curve further confirmed the value of the proportions of CD3^+^ and CD8^+^ T cells in predicting the severity of COVID-19.

As a retrospective study, this study has some notable limitations. Firstly, due to the requirements of the government to treat COVID-19 patients according to disease grades, our institute only treated mild and severe patients, while critically ill patients were transferred to other designated hospitals. This study was performed in a single centre and only included patients with mild and severe types of COVID-19. Secondly, since the data generation was clinically driven and not systematic, we did not include other markers that have been associated with the outcomes of viral infectious diseases, such as D-dimer. Several patients did not receive procalcitonin and sputum pathogenic microbe detection tests due to overwhelmed medical resources.

## Conclusion

Metabolic disturbances and immune-inflammatory dysfunction were found in patients with COVID-19. Progressive lymphopenia, hypoproteinaemia, hypoalbuminemia, low high-density lipoproteinemia, and decreased level of ApoA1, CD3^+^T%, and CD8^+^T% indicate worse outcomes and severe type of COVID-19.

## Data Availability

No data available in this manuscript.

## Notes

### Contributors

S-KN and X-QZ contributed equally to this article. S-KN and ZZ conceptualized the paper. S-KN analysed the data, with input from KZ, X-QZ and Z-HZ. S-KN and Z-TZ wrote the initial draft with all authors providing critical feedback and edits to subsequent revisions. All authors approved the final draft of the manuscript. ZZ is the guarantor. The corresponding author attests that all listed authors meet authorship criteria and that no others meeting the criteria have been omitted.

## Acknowledgements

We would like to thank Editage (*www.editage.cn*) for English language editing.

## Funding

This work was supported by the emergency science and technology projects of COVID-19 of Hubei Province (No.2020FCA005).

## Conflict of interests

All author had no conflict of interests to declare. No other relationships or activities that could appear to have influenced the submitted work. This study has never previously been presented in any meetings.

## Ethical approval

This study was approved by the Ethics Committee of Renmin Hospital of Wuhan University (WDRY2020-K100). Requirement for written informed consent was waived by Renmin Hospital of Wuhan University.

## Notes

### Competing Interest Statement

The authors have declared no competing interest.

